# A rising burden of adolescents obesity of age group 13-17 years among tribal population of central India: a community-based exploratory study

**DOI:** 10.1101/2021.01.12.20249097

**Authors:** Nikhil Shukla, Prabal Kumar Chourasia, Somit Kumar Jain, Aravind Lathika Rajendrakumar, Anand Thakarakkattil Narayanan Nair, Charvi Nangia, Mehul Kumar Chourasia

**Affiliations:** Sri Sathya Sai Sanjeevani Hospital, Raipur, Chhattisgarh India; Mary Washington Medical Group, Fredericksburg, VA, USA; Prestige Dental Care, Raipur, Chhattisgarh India; School of Medicine, University of Dundee, Dundee, UK

**Keywords:** Adolescent, Body Mass Index, Obesity, Tribals, India

## Abstract

**Background:** Upsurge of adolescent obesity is an upcoming national public health concern. Obese adolescents are at significant risk of becoming obese adults and its co-morbidities. This study estimates the prevalence of adolescent obesity and explore the potential determinants among young adults residing in tribal populated villages of Chhattisgarh, India.

**Methods:** A community-based nutritional survey was carried out among adolescent of the age group of 13-17 years.

**Results:** Among 1,296 participants, 23.4 % of young adults were either overweight or obese. Higher family earnings (Odds ratio [OR], 2.79, 95% confidence interval [CI] 1.29-6.38), Skipping breakfast (3.09, 1.11-8.30), Television viewing > 2 hours/ day (2.16, 1.3-6.2), Energy intake (2.98, 1.19-15.6), significantly increased the risk of adolescent obesity.

**Conclusion:** Prevalence of adolescence obesity among the tribes is alarming and needs to be tackled with health system measures. Future research may require assessing the trajectory of obesity and related comorbidities in a tribal population.

## Introduction

Obesity is an established risk factor for many chronic diseases such as diabetes, hypertension, and premature mortality. Previous studies and systematic reviews have suggested that obese adolescents are at significant risk of becoming obese adults and a high economic cost associated with adult obesity and its comorbidities^1,2,3^. There is still a social stigma surrounding obesity and there has been a push from the scientific community to formally declare obesity as a disease.^4^ Such an approach aimed to manage obesity in a better way and make it socially acceptable to generate tangible health benefits. Obesity is a global health problem as illustrated by the fact that 340 million in 5-19 year age group in 2016 worldwide were obese.^5^ This indicates that obesity is a global pandemic and not just confined to developed nations. Furthermore, overweight or obese adolescents have a propensity to become heavier in their adulthood.^6^

It is estimated that 135 million in India is suffering from some degree of obesity most of which is driven by sedentary lifestyles promoted by socioeconomic and epidemiological transition.^7^ India is an ethnically diverse country and there is heterogeneity in adiposity between population subgroups.^8^ There is a paucity of data regarding the prevalence of obesity and its risk factors among the socially vulnerable groups such as the tribal community. The main objective of this study is to estimate the prevalence of adolescent obesity and explore the potential determinants among young adults residing in tribal populated villages of Chhattisgarh, India.

## Materials and Methods

A community-based nutritional survey was carried out as a part of health awareness camp in a tribal populated sub-district of Raipur, Chhattisgarh from December 2017 to June 2018. The complete household list was obtained from the panchayat office. Adolescent of the age group of 13-17 years was selected via systematic random sampling. Written consent of the Individual and head of the family was obtained before administration of the interview. Information from previous day diet intake history and “modified version of the international physical activity questionnaire” ^9^ was collected. For the diet history, participants were asked about all the food and drinks consumed during the previous day. Their dietary intake information was taken in household measurements and was converted to grams by using standard reference tables. BMI (kg/m^2^) was calculated as weight (in kilograms) divided by height^2^ (in meters). BMI–for–age above +2SD of WHO 2007 growth standards, were considered as obese.^10^ For the Physical activity pattern, Total physical activity pattern (in MET-week) for a typical week was assessed. It includes three activities: vigorous, moderate and walking. Participants were categorized into three groups; physically less active, moderate, and high active based on IPAQ cut off values. Details of the sample size calculation, energy intake, an annual household earning and physical activity were given in supplementary methods.

The statistical analysis was carried out by using the STATA software. Univariate and multivariate logistic regression analysis was performed to analyse the association between potential categorical variables. The odds ratio (OR) and 95% confidence interval (CI) were also assessed and computed for each significant categorical variable using binary logistic regression. A p-value < 0.05 was considered as statistically significant. The goodness of fit of the final model was assessed using the Hosmer-Lemeshow technique, (p-value >0.05).

Present study as part of health awareness camp was discussed and approved by the Institutional Ethical Review board of e-Aarogyam Gyan and Health foundation, India (IRC/017/09.01) and results were used to plan for the health education intervention content in the area specific to the need of the tribal population.

## Results

We analysed data from 1,296 adolescents and among them, 23.4 % were either overweight or obese (Table 1). We had an almost equal proportion of participants among both females and males as well as for all ages (13-17 years). There were no notable differences between gender and underweight, obesity and overweight (suppl fig 1). Around 18% and 8% of the participants were obese and underweight, respectively. Table 2 illustrates that higher family earnings (Odds ratio [OR], 2.79, 95% confidence interval [CI] 1.29-6.38), Skipping breakfast (3.09, 1.11-8.30), Consumption of fruits < 4 days per week (2.18, 1.02-4.67), Television viewing > 2 hours/ day (2.16, 1.3-6.2), Energy intake (2.98, 1.19-15.6), significantly increased the risk of obesity, whereas increased physical activity (5.34, 1.68-15.58) decreased the odds of getting obese. Also, Irregular menstruation (3.34, 1.27-12.10) was noted among obese girls.

**Table 1:**
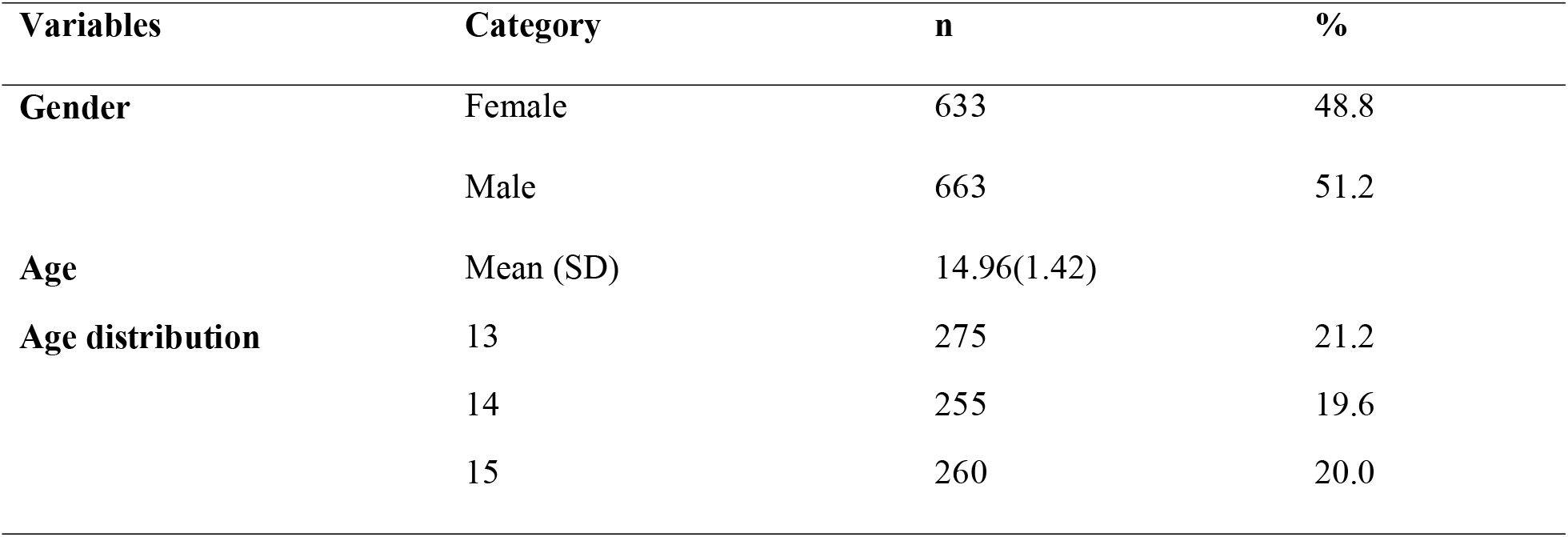
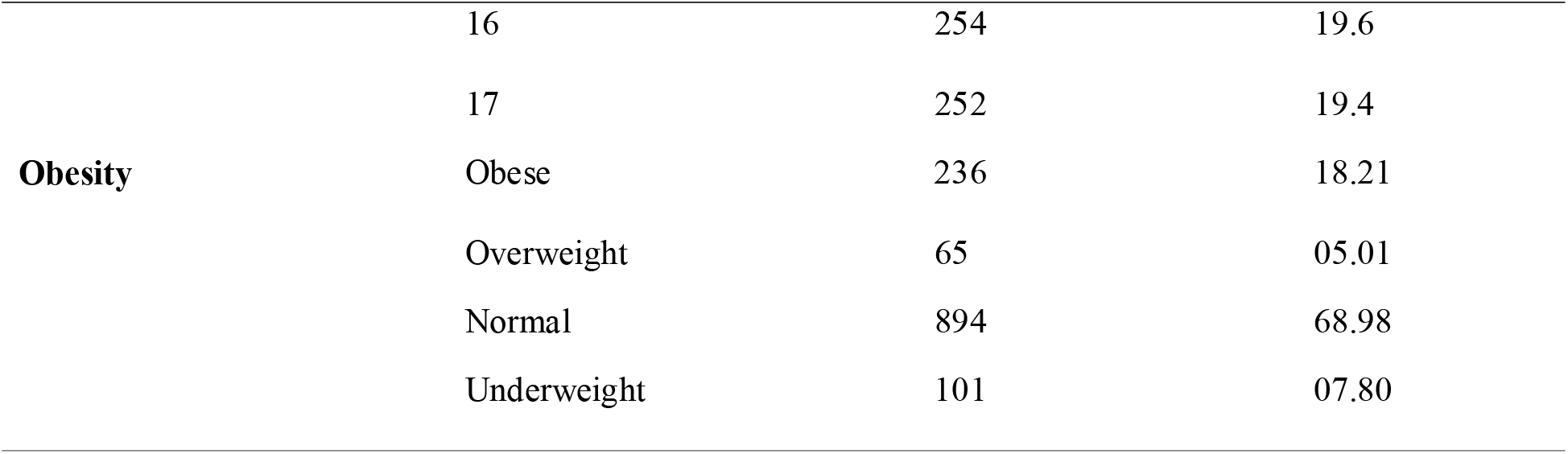
Descriptive Statistics of study population (n=1,296)

**Table 2.** Potential risk factors associated with adolescent obesity (n=1,296)

| **Study Variables** | **Univariate**<br>**OR** | **P value** | **Multivariate**<br>**OR\*** | **95% CI** | **P-value** |
| --- | --- | --- | --- | --- | --- |
| Higher family earnings | 3.42 | 0.001* | 2.79 | 1.29-6.38 | <0.01* |
| Skipping breakfast | 3.19 | <0.001* | 3.09 | 1.11-8.30 | <0.01* |
| Consumption of fruits < 4<br>days per week | 2.93 | 0.01* | 2.18 | 1.02-4.67 | 0.04* |
| Television viewing > 2<br>hours/ day | 3.3 | 0.001* | 2.16 | 1.3-6.2 | <0.01* |
| Energy intake | 6.7 | 0.001* | 2.8 | 1-3.4 | 0.05 |

Also adjusted for Age and sex; \* Statistically significant

## Discussion and Conclusion

This is one of the first studies which highlights the epidemiological transition and dual burden of both undernutrition and obesity among a tribal community in the central part of India. Tribal population in India are disadvantaged both socially and economically due to several historical reasons. India has a sizeable proportion of the tribal community in its population particularly in eastern central India and translates to 8% of the Indian population in 2011.^11,12^ The major finding of the current study is the high burden of obesity among adolescents of the indigenous population, 18.2%. with another 5% being overweight. This shows that the obesity burden of tribal in Chhattisgarh is much higher than those reported in tribal and closer to the burden in Dhodias (23.8%).^13^ The reported prevalence of undernutrition 7.8% is also much less than those previously reported.^13^ Therefore, while the reduction in undernutrition is encouraging, but a steep rise in obesity is a cause of concern. It should be also noted that tribal people from the same region were developed diabetes and other chronic diseases even at normal BMI.^14^

The major limitation of the study is that study was carried out in a subsection of the tribal population and therefore may not be generalizable to the tribal community as a whole. Some of the obesity risk factors and obesity indices such as Waist-Hip ratio were not captured in this study.

In conclusion, rising obesity in the tribal community is alarming and needs to be tackled with urgent policy and health system measures. Future research may explore obesity in detail with multiple obesity indices and trajectory of obesity and related comorbidities in a tribal population.

## Authors’ contributions

NS, and PKC designed the study. MKC, ALR, ATN and CN drafted the manuscript. MKC, and ALR have done the literature search and prepared the study protocol. NS and SKJ supervised, collected, and cleaned the data. MKC, ATN, and ALR analysed the data. NS, PKC, SKJ, ALR, ATN, CN, and MKC reviewed and helped to write the manuscript. All authors have given the intellectual input to the study. All authors read and approved the final manuscript.

## Acknowledgement

We express our gratitude to the study participants for their enthusiastic cooperation with this survey. We would also like to acknowledge e-Aarogyam Gyan and Health foundation, India for their valuable support. We would also like to extend our gratitude to Dr Anand Verma, and Dr Somit Kumar Jain for their sincere efforts in data collection and logistics management.

## Data availability

‘The datasets used and analyzed during the current study are available from the corresponding author on reasonable request.’

