## Supplementary material for "A rising burden of adolescents obesity of age group 13-17 years among tribal population of central India: a community-based exploratory study": Methods

**SUPPLEMENTARY FILE**

**Methods**

**Sample size calculation^1^**

Study sample size (n) = 1,140 [ N = 100,000, d = 2.5 % , p = 25% ]

Sample size (n) was calculated for the cross-sectional survey using the following formula:

n = N*X / (X + N – 1); where, X = Z_α/2_^2^ ­*p*(1-p) / d^2^,

Z_α/2_ = (95% confidence level (CI), α = 0.05) = 1.96; d = margin of error; p = assumed sample proportion, N = total population size

**An annual household earning and Socio-economic status**

Most of the tribal community in the study area live in a similar housing type and education level ranges from primary to lower middle class. Hence, only monthly earning has been used for the current analysis. To classify the annual family earning, SES scale developed by Tiwari et al have been used^2^.

**Energy Intake calculation**

Semi-quantitative food frequency questionnaire (FFQ) was used for energy intake.^3^

**Physical activity level**

A short version of PAQ was used for the classification of physical activity.^4^

Physical activity levels were categorized based on the number of minutes of participation in either moderate-intensity and/or vigorous-intensity activity/ week.

Low physical activity = < 75 min of vigorous-intensity activity per week or less than 150 min of moderate-intensity physical activity/week

Medium physical activity = 150–300 min of moderate-intensity activity or 75–150 min of vigorous-intensity physical activity/week

High physical activity level = > 300 min of moderate-intensity physical activity/ week.
