## Supplemental Figure 1 for "A rising burden of adolescents obesity of age group 13-17 years among tribal population of central India: a community-based exploratory study"

Supplementary Figure 1. Sex disaggregated proportion of obesity, overweight and underweight among tribal adolescents (13-17 years)

| Gender | Percentage |
| --- | --- |
| Male obese | 18.9 |
| Male overweight | 4.7 |
| Male underweight | 8.1 |
| Female obese | 17.5 |
| Female overweight | 5.4 |
| Female underweight | 7.4 |
